# Urinalysis, but not blood biochemistry, detects the early renal-impairment in patients with COVID-19

**DOI:** 10.1101/2020.04.03.20051722

**Authors:** Haifeng Zhou, Zili Zhang, Heng Fan, Junyi Li, Mingyue Li, Yalan Dong, Weina Guo, Lan Lin, Zhenyu Kang, Ting Yu, Chunxia Tian, Yang Gui, Renjie Qin, Haijun Wang, Shanshan Luo, Desheng Hu

## Abstract

**Background:** In December 2019, a novel coronavirus (SARS-CoV-2) caused infectious disease, termed COVID-19, outbroke in Wuhan, China. COVID-19 patients manifested as lung injury with complications in other organs, such as liver, heart, gastrointestinal tract, especially for severe cases. However, whether COVID-19 causes significant acute kidney injury (AKI) remained controversial.

**Methods:** We retrospectively analyzed the clinical characteristics, urine and blood routine tests and other laboratory parameters of hospitalized COVID-19 patients in Wuhan Union Hospital.

**Findings:** 178 patients, admitted to Wuhan Union hospital from February 02 to February 29, 2020, were included in this study. No patient (0 [0%]) presented increased serum creatinine (Scr), and 5 (2.8%) patients showed increased blood urea nitrogen (BUN), indicating few cases with “kidney dysfunction”. However,for patients (83) with no history of kidney disease who received routine urine test upon hospitalization, 45 (54.2%) patients displayed abnormality in urinalysis, such as proteinuria, hematuria and leukocyturia, while none of the patients was recorded to have acute kidney injury (AKI) throughout the study. Meanwhile, the patients with abnormal urinalysis usually had worse disease progression reflecting by laboratory parameters presentations, including markers of liver injury, inflammation, and coagulation.

**Conclusion:** Many patients manifested by abnormal urinalysis on admission, including proteinuria or hematuria. Our results revealed that urinalysis is better in unveiling potential kidney impairment of COVID-19 patients than blood chemistry test and urinalysis could be used to reflect and predict the disease severity. We therefore recommend pay more attention in urinalysis and kidney impairment in COVID-19 patients.

## Introduction

A novel coronavirus, named SARS-CoV-2, caused pneumonia outbreak in December 2019 in Wuhan, China, and quickly spread across the country. Until March 16, 2020, there have been 16,7511 confirmed cases of COVID-19 and 6606 deaths worldwide (announced by WHO). Despite most cases were from China from the beginning, local outbreak has occurred in other countries recently, such as Japan, South Korea, Iran and Italy. Epidemiological investigations demonstrated that elderly men were susceptible to the disease and more likely to develop into severe or critically ill stage(1-3). Furthermore, comorbidities, such as diabetes, hypertension, cardiovascular disease and cancers, are risk factors for COVID-19(1, 2). Though most of the COVID-19 patients usually initiated with flu-like symptoms, some of them underwent rapid deterioration and even die suddenly(4, 5). Emergency measures including quarantine, traffic control and suspension of public activities are implemented to limit the epidemic. However, there is no effective treatment against COVID-19 and the presence of asymptomatic carriers and the possibility of relapse impose additional challenges for COVID-19 prevention(6, 7).

Genomic sequence analysis indicated that SARS-CoV-2 is 96% identical to bat coronavirus and has 75%-80% similarity to severe acute respiratory syndrome coronavirus (SARS-CoV) (8-10). SARS-CoV-2 uses angiotensin converting enzyme II (ACE2) as cell entry receptor, thus, patients with COVID-19 could develop multiple organ or tissue injuries(11-14). It is reported that the patients with SARS-CoV-2 infection show elevated levels of alanine aminotransferase (ALT), aspartate aminotransferase (AST) and γ□ glutamyl transferase (GGT), indicating liver and myocardial damage(1, 3, 15-17). Yet the existence of COVID-19 induced kidney impairment or dysfunction remain controversial. A recent study reported that 29% of the 52 patients with severe SARS-CoV-2 pneumonia were complicated with acute kidney injury (AKI)(4). Another study among 138 hospitalized patients with confirmed COVID-19 revealed that 5 patients (3.6%) developed AKI and 3 of them ended up in the ICU. Yan et al. demonstrated that among the 59 COVID-19 patients with kidney impairment, 63% exhibited proteinuria, 19% displayed elevated level of plasma creatinine and 27% showed increased level of urea nitrogen(18). A study from Xu’s group reported that 5.1% (36/701) patients were identified as AKI(19). In addition, Huang et al revealed that AKI occurred in 7% (3/41) patients(2). On the contrary, a study with 1099 confirmed COVID-19 cases indicated that only 6 patients (0.5%) developed AKI(17). Another study conducted by Zhou et al revealed that only 4% (8/186) patients had increased level of Scr(1). Gong et al. reported that SARS-CoV-2 infection could neither cause obvious AKI nor aggravate CRF in the COVID-19 patients(20). Thus, as with other known extrapulmonary organ injury, the existence of AKI deserves to be disclosed in COVID-19 patients, which is pivotal in the disease diagnosis, treatment and prognosis.

The current study retrospectively analyzed the clinical data of 178 hospitalized COVID-19-confirmed patients in Wuhan Union Hospital and proposes to provide additional evidence on the incidence of kidney injury in COVID-19 patients and address a better indicator for the potential kidney impairment.

## Method

### Study design

178 patients with confirmed COVID-19 in Wuhan Union hospital, Tongji Medical College, Huazhong University of Science and Technology, from February 02 to February 29, 2020, were enrolled in this retrospective study. All patients in this study were diagnosed according to the guidance (fifth edition) published by Chinese National Health Commission. The patients with a history of kidney diseases were excluded from this study, and the flow chart was shown in **Figure 1**. And the clinical outcomes were monitored up to February 29, 2020. This study was approved by the Institutional Ethics Board of Wuhan Union Hospital of Tongji Medical College, Huazhong University of Science and Technology (No. Union Hospital-0093). Written informed consent was waived by the Ethics Commission of the designated hospital for the emerging infectious diseases.

**Figure 1.**
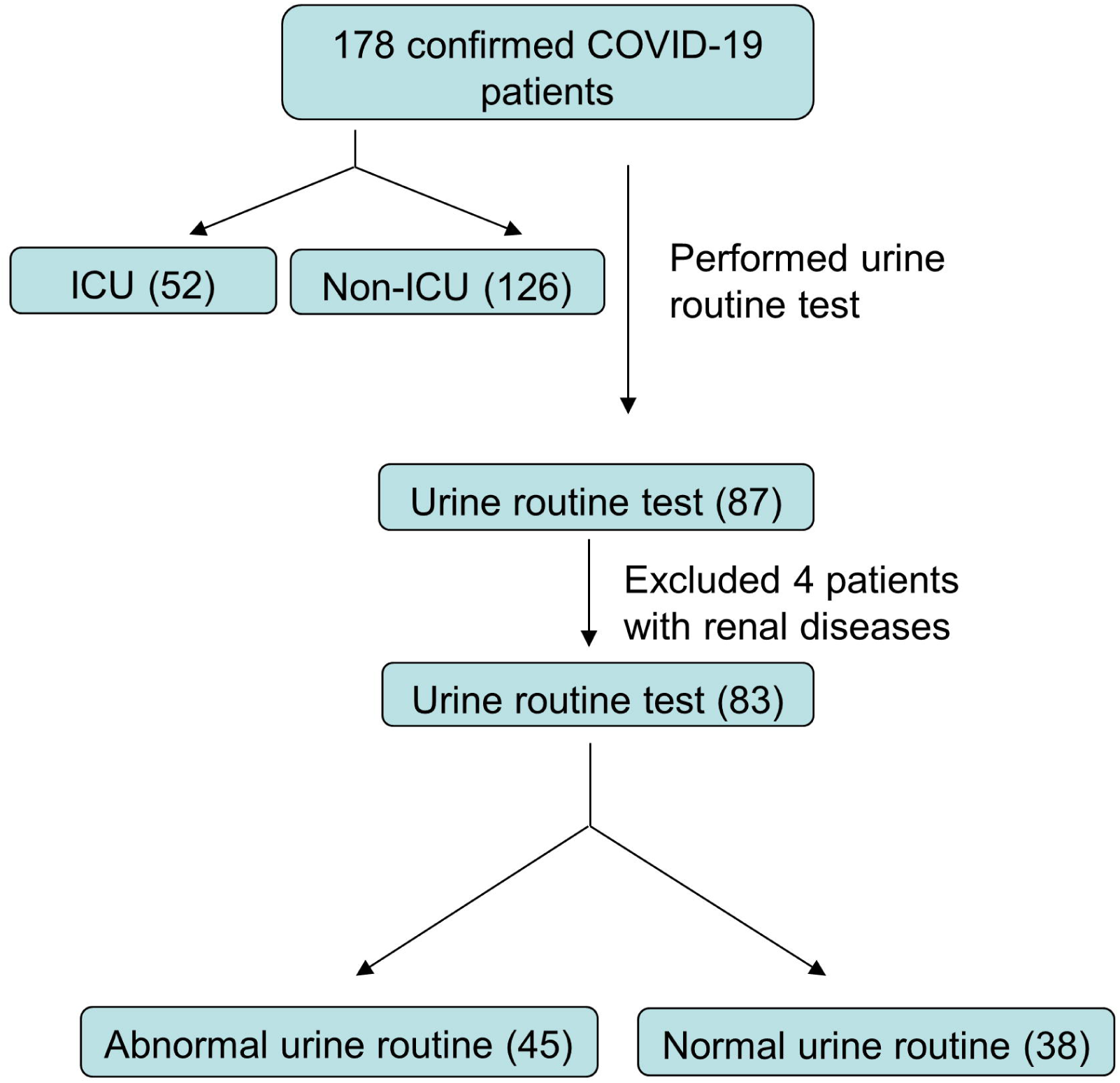
The flow chart of COVID-19 patients inclusion procedure.

### Data collection

Demographic data, symptoms and vital signs, laboratory examinations were retrieved from electronic medical records for this study. The laboratory examinations contain blood routine test, blood biochemical tests (liver function, renal function, lipids and glucose), blood coagulation index, lymphocyte subsets and cytokines analysis, urine routine test, C-reactive protein (CRP), erythrocyte sedimentation (ESR), serum ferritin (SF), etc. Estimated glomerular filtration rate (eGFR) was calculated with Chronic Kidney Disease Epidemiology Collaboration (CKD-EPI) equation(21). Patients were daily assessed the presence of AKI or renal failure using strictly criteria based on the 2011 kidney disease: improved global outcomes (KDIGO) for AKI. All the laboratory measures were performed at the department of clinical laboratory in Union Hospital and the normal range of these measures were given by them. The threshold of Scr was 54-133 (μmol/L) for male and 44-106 (μmol/L) for female, BUN was 2.9-8.2 (mmol/L), serum uric acid was 208-428 (μmol/L) for male and 155-357 (μmol/L) for female, as well as Cystatin C was 0.63-1.25 (mg/L) for male and 0.54-1.15 (mg/L) for female. Proteinuria was defined as positive in urine protein (semiquantitative, +-/+/++/+++) and hematuria was recorded with elevated urine erythrocyte quantification (>17/ul) as well as positive in semiquantitative urinalysis (+-/+/++/+++). All the parameters were analyzed using extreme values, except for urinalysis upon first examination on admission due to limited data.

### Statistical analysis

The categorical variables were summarized as counts or percentages, and continuous variables were expressed as mean ± SD or medians and interquartile ranges (IQR) as appropriate. Categorical variables were compared using the χ^2^ test; the Fisher exact test was used when the data were limited. Continuous variables were compared using independent group t test when the data were normally distributed; otherwise, the Mann-Whitney test was used. All statistical analyses were performed using SPSS 20.0 software, and P <0.05 was regarded as a significant difference.

## Result

### COVID-19 patients were seldom diagnosed with AKI

A total of 178 patients with laboratory confirmed SARS-CoV-2 infection were enrolled in this study, among which 52 patients were from ICU and 126 patients were from ordinary isolation ward. As shown in **Table 1**, the median age of patients in ICU group was 56.5 years (IQR, 40.50-65.5), which was significantly higher than that of patients in non-ICU group (45, IQR [32.75-57.00]). The proportion of males in ICU group was higher than that of non-ICU group (61.5% vs. 31.7%). More patients in ICU group rather than non-ICU group manifested high body temperature (T>38 □) (75% vs. 56.3%). Additionally, some underlying diseases were more common in patients of ICU, such as hypertension (28.8% vs. 11.1%), diabetes (17.3% vs. 7.9%), cardiovascular insults (13.5% vs. 2.4%), digestive system (7.7% vs. 4.8%) and respiratory system disease (7.7% vs. 4.8%). These data were consistent with recent reports related to COVID-19 that elder men with comorbidities are more likely to be infected and develop into severe cases and even die due to weaker immune function.

**Table 1.** Basic information and blood indicators related to kidney injury

|  | Total<br>(N=178) | ICU<br>(n=52) | Non-ICU<br>(n=126) | <sup>a</sup>P Value |
| --- | --- | --- | --- | --- |
| **Age (Years)** | 47.0 (35.0-61.0) | 56.50 (40.5-65.5) | 45.0 (32.8-57.0) | <0.001 |
| **Sex** |  |  |  |  |
| male | 72 (40.4) | 32 (61.5) | 40 (31.7) | <0.001 |
| female | 106 (59.6) | 20 (38.5) | 86 (68.3) | <0.001 |
| **Basic disease** |  |  |  |  |
| hypertension | 29 (16.3) | 15 (28.8) | 14 (11.1) | 0.004 |
| hyperlipidemia | 3 (1.7) | 1 (1.9) | 2 (1.6) | >0.999 |
| diabetes | 19 (10.7) | 9 (17.3) | 10 (7.9) | 0.066 |
| digestive system | 10 (5.6) | 4 (7.7) | 6 (4.8) | 0.481 |
| respiratory system | 10 (5.6) | 4 (7.7) | 6 (4.8) | 0.481 |
| cardiovascular | 10 (5.6) | 7(13.5) | 3 (2.4) | 0.007 |
| cancer | 8 (4.5) | 1 (1.9) | 7 (5.6) | 0.440 |
| urinary system | 4 (2.2) | 2 (3.8) | 2 (1.6) | >0.999 |
| Others | 23 (12.9) | 5 (9.6) | 18 (14.3) | 0.398 |
| non | 50 (30.3) | 14 (30.8) | 36 (30.2) | 0.936 |
| **Body temperature (max.)**<br>**(□)** | 38.5 (37.8-39.0) | 39 (38.1-39.1) | 38.3 (37.7-39.0) | 0.005 |
| >38 | 110 (61.8) | 39 (75) | 71 (56.3) | 0.020 |
| **Scr (μmol/L)** | 65.2 (56.8-74.8) | 71.0 (55.8-89.4) | 65.3 (56.5-74.3) | 0.067 |
| increase | 0 | 0 | 0 | >0.999 |
| **BUN (mmol/L)** | 3.81 (2.83-4.54) | 4.14 (3.33-4.82) | 3.72 (2.76-4.57) | 0.078 |
| increase | 5 (2.8) | 3 (5.8) | 2 (1.6) | 0.150 |
| **eGFR (ml/min)** | 214.6 (308.05) | 93.7 (78.1-108.0) | 102.5 (90.7-113.1) | 0.014 |
| decrease | 42 (23.6) | 19 (36.5) | 23 (18.3) | 0.009 |
| **Cystatin C (mg/L)** | 0.77 (0.68-0.86) | 0.81 (0.73-0.88) | 0.77 (0.68-0.87) | 0.155 |
| increase | 3 (1.7) | 1 (1.9) | 2 (1.6) | >0.999 |

**Note:** Data are shown as median (IQR) or n (%) as appropriate. ICU=intensive care unit, Scr=serum creatinine, BUN=blood urea nitrogen, eGFR=estimated glomerular filtration rate, IQR=interquartile range. <sup>a</sup> P values indicate differences between ICU and Non-ICU patients. P <0.05 was considered statistically significant.

Previous studies showed that many patients with SARS-CoV-2 infection usually had increased level of Scr, BUN and even AKI, especially those severe patients. Unexpectedly, these clinical manifestations, however, didn’t appear in our data. Patients in both ICU group and non-ICU group displayed normal Scr and cystatin C level. Of all cases, although 23.6% patients showed decreased estimated glomerular filtration rate (eGFR) compared to normal level, only 2.8% patients were recorded with elevated level of BUN. Consistent with these results, none of patients develop acute kidney injury or acute renal failure during whole hospitalization regardless of entry into ICU. Taken together, our data supported that there was no significant kidney dysfunction among COVID-19 patients.

### Potential kidney impairment reflected by routine urine test

Based on the fact that Scr and BUN may not be affected owing to strong compensatory adaption of kidney, urine examination results were collected to further evaluate the kidney impairment in detail. Not every COVID-19 patient received regular urine routine test. Only 87 of 178 enrolled patients underwent urine test on admission, among whom 4 patients were documented with history of kidney diseases: diabetic nephropathy, kidney stones, multiple renal cystic disease and kidney carcinoma. And they were excluded in our cohort. The remaining 83 patients were selected for further analysis as illustrated in **Figure 1**, with a participation rate of 46.6%. Of the 83 patients, no one reported related symptoms or was diagnosed as urinary system infection, urethral injury or other urinary system diseases. Unexpectedly, 54.2% (45/83) patients showed abnormal urine test results, including proteinuria, hematuria, leukocyturia **(Table 2)**. Specifically, 34.9% cases had positive urine protein, with 15.7% for “+-”, 16.9% for “+” and 2.4% for “++” **(Figure 2A)**. At the same time, 31.3% patients presented with hematuria, including 2.4% for “+-”, 16.9% for “+”, 8.4% for “++” and 3.6% for “+++” **(Figure 2A)**. In addition, the patients in ICU exhibited more severe urinalysis results compared with non-ICU patients, such as proteinuria (+, 31.6% vs. 12.5%; ++,10.5% vs. 0) and hematuria (+, 26.3% vs. 18.8%; ++, 21.1% vs. 4.7%; +++, 5.3% vs. 3.1%) (P<0.05 for all) **(Figure 2B, C)**. These results indicated prevalence of kidney impairment among COVID-19 patients even without significant changes of filtration function.

**Table 2.** Abnormal urinalysis results in COVID-19

|  | Total<br>(N=83) | ICU<br>(n=19) | Non-ICU (n=64) | <sup>a</sup>P Value |
| --- | --- | --- | --- | --- |
| **Abnormal urine routine** | 45 (54.2) | 15 (78.9) | 30 (46.9) | 0.014 |
| **Proteinuria** |  |  |  |  |
| positive | 29 (34.9) | 11 (57.9) | 18 (28.1) | 0.017 |
| **Hematuria** |  |  |  |  |
| positive | 24 (28.9) | 10 (52.6) | 14 (21.9) | 0.009 |
| **Leucocyturia** |  |  |  |  |
| positive | 14 (16.9) | 2 (10.5) | 12 (18.8) | 0.506 |
| **Urine glucose** |  |  |  |  |
| positive | 0 (0) | 0 (0) | 0 (0) | >0.999 |
| **Urine urothelial cell** |  |  |  |  |
| positive | 10 (12) | 1 (5.3) | 9 (14.1) | 0.441 |

**Note:** Data are shown as n (%). COVID-19=novel coronavirus disease 2019, ICU=intensive care unit. <sup>a</sup> P values indicate differences between AU and NU patients. *P* <0.05 was considered statistically significant.

**Figure 2.**
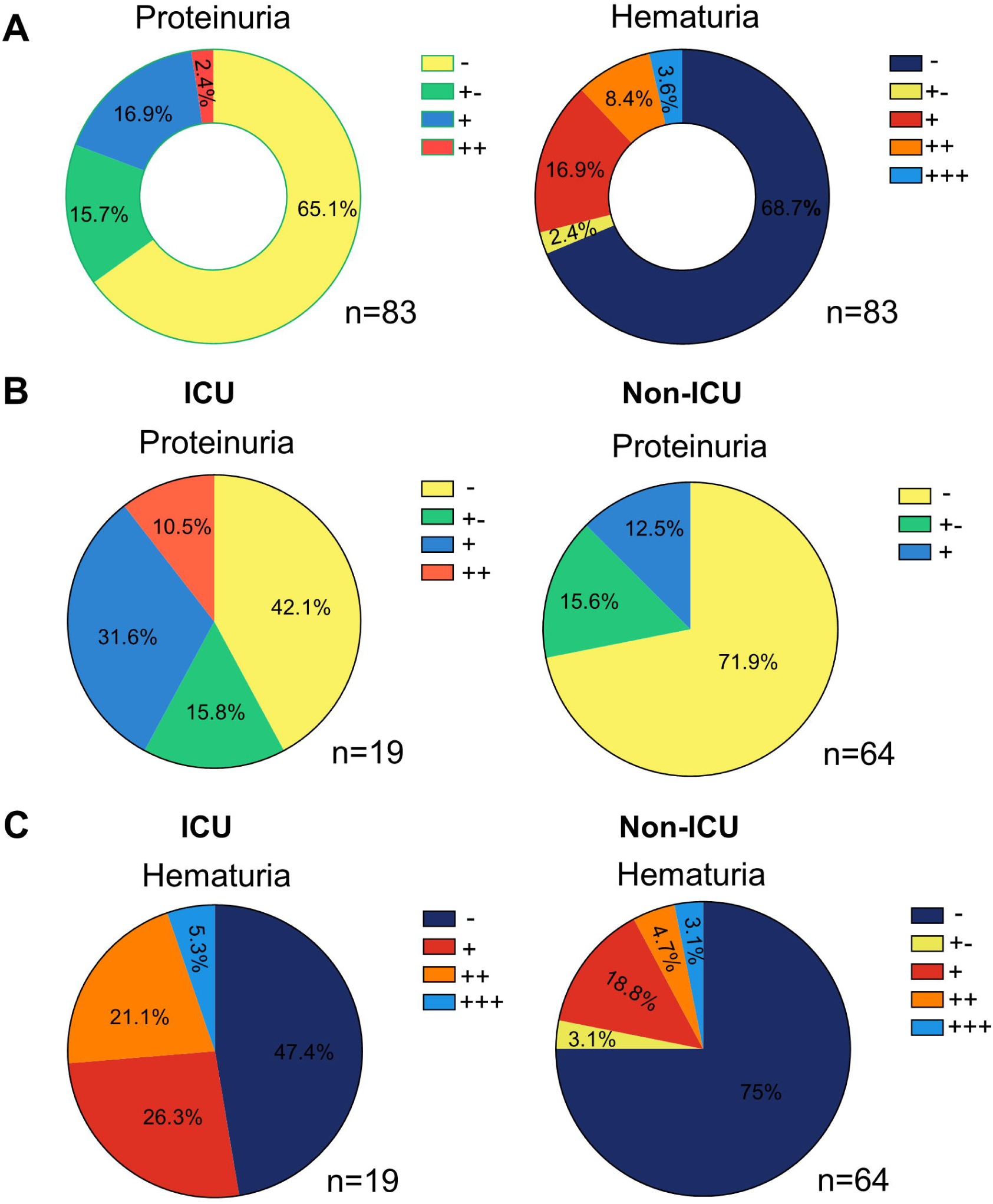
Pie chart illustrating COVID-19 patients exhibited abnormal urinalysis, including. **proteinuria and hematuria. (A)** Analysis of all the COVID-19 patients with urine routine test. **(B)** Analysis of proteinuria between ICU and non-ICU. **(C)** analysis of hematuria between ICU and non-ICU.

### Kidney impairment were caused by direct virus invading

Given that proteinuria and hematuria were two prominent signs of kidney impairment, 83 patients were allocated to groups with abnormal urinalysis (AU) or normal urinalysis (NU) based on urinalysis. It is also possible that the kidney impairment is due to drug nephrotoxicity. The medications of patients before admission included antibiotics (Moxifloxacin, Azithromycin, Amoxicillin) (30.1%), antiviral drugs (Oseltamivir, Lopinavir, Arbidol) (33.8%) and Chinese patent medicine (Lianhuaqingwen capsule, Isatis root granule) (10.8%). However, there was no statistical difference in prehospital treatment between AU and NU group **(Table 3)**. This data supported that the kidney impairment was caused by SARS-CoV-2 infection rather than treatment agents.

**Table 3.** Prehospital medications between AU and NU groups

| Medications before admission | Total (N=83) | AU (n=45) | NU (n=38) | <sup>a</sup>P Value |
| --- | --- | --- | --- | --- |
| Antibiotic | 25 (30.1) | 11 (35.5) | 14 (26.9) | 0.411 |
| Oseltamivir / Lopinavir | 12 (14.5) | 7 (22.6) | 5 (9.6) | 0.119 |
| Arbidol | 16 (19.3) | 9 (29.0) | 7 (13.5) | 0.082 |
| Chinese Patent Medicine | 9 (10.8) | 3 (9.7) | 6 (11.5) | >0.999 |

**Note:** Data are shown as n (%). AU=abnormal urinalysis group, NU=normal urinalysis group. <sup>a</sup> P values indicate differences between AU and NU patients. Chinese Patent Medicine consists of Lianhuaqingwen capsule and Isatis root granule. *P* <0.05 was considered statistically significant.

### Urinalysis abnormality correlates with severity of COVID-19

Subsequently, in order to explore whether urinalysis correlates with disease severity, we compared the differences in other laboratory parameters between the AU and NU group. Firstly, we found that there was no difference in eGFR, Scr, BUN and serum uric acid between the two groups. Although the patients with AU had higher level of cystatin C, all the values were generally within normal range **(Figure 3A)**. Next, as shown in **Figure 3B**, the patients in AU group demonstrated remarkably higher level of liver injury related indicators, such as ALT, AST, GGT, α-hydroxybutyrate dehydrogenase (HBDH), lactic dehydrogenase (LDH), alkaline phosphatase (ALP) and leucine aminopeptidase (LAP). And both serum prealbumin level and albumin-globulin ratio (A/G), markers related to liver synthetic capability and inflammatory state, were lower in AU group than in NU group, indicating possible liver inflammation. These data revealed that the patients with abnormal urinalysis might be subjected to liver damage. In addition, the patients in AU group usually presented with higher levels of inflammation related markers, including C-reactive protein (CRP), erythrocyte sedimentation rate (ESR), IL-6, serum ferritin (SF) and serum amyloid A (SAA) **(Figure 3C)**. However, no significant differences were observed in major lymphocytes subpopulations, such as CD4^+^ T cells, CD8^+^ T cells, CD4/CD8, B cells and NK cells **(Figure 3D)**. What’s more, absolute count of peripheral lymphocyte of AU patients was significantly lower than that of NU patients, white blood cells (WBC) and neutrophils count were usually higher in patients in AU group **(Figure 3E)**. There was no significant difference between the two groups in platelet, red blood cells (RBC) and hemoglobin **(Figure 3E)**. As for coagulation profile, the AU patients had statistically higher levels of serum fibrinogen (FIB) and D-dimer than NU patients, indicating the worse coagulation function **(Figure 3F)**. Collectively, these results indicated that the urinalysis abnormality correlates with severity of COVID-19, and urine routine test may be a good method for predicting the disease progression in COVID-19 patients.

**Figure 3.**
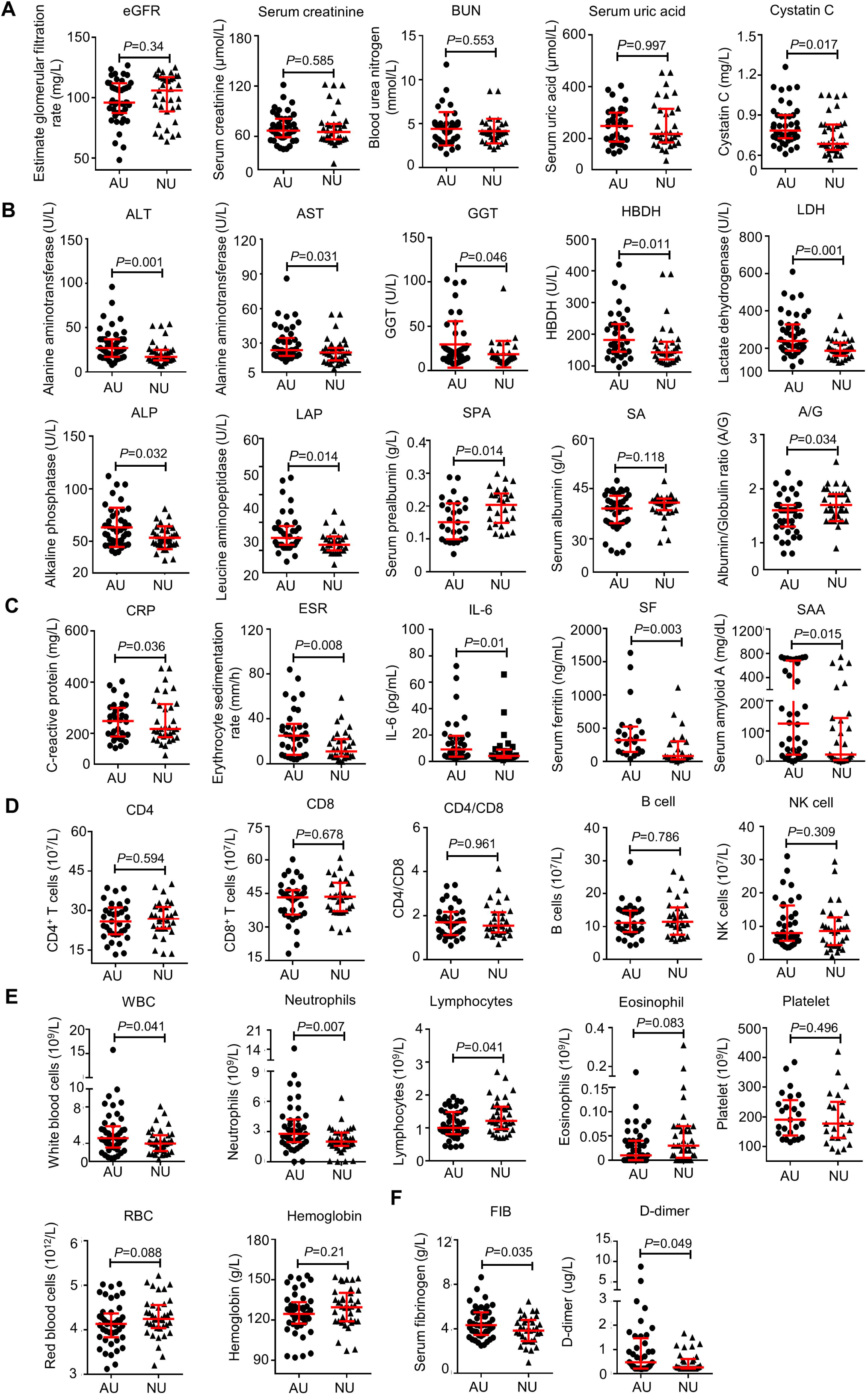
The comparison of laboratory parameters between abnormal urinalysis group and normal urinalysis group. Analysis of renal function **(A)**, analysis of liver function **(B)**, inflammation **(C)**, lymphocytes subpopulations **(D)**, blood routine **(E)** and coagulation function **(F)**. Abbreviation: GGT, γ*-*glutamyl transpeptidase; HBDH: α-hydroxybutyrate dehydrogenase.

## Discussion

Here, we described the kidney impairment related manifestations among COVID-19 patients. Our results suggest kidney impairment are common in patients with SARS-CoV-2 infection, however, acute kidney injury occurs infrequently and primary symptoms were abnormal urinalysis, including proteinuria, hematuria, and leucocyturia. At the same time, urine routine test can be used as a good method that may be neglected to predict disease severity.

Several papers have demonstrated the renal impairment in COVID-19 patients. However, the proportion of increased Scr or incidence of AKI after SARS-CoV-2 infection was quite different among these studies. Some researches supported that SARS-CoV-2 infection cause significant renal impairment, however, some held an opposite view. We carefully read the relevant articles and found that the paradox may result from: (1) the difference in severity of illness; (2) the different scale of patient sampling. Further investigations are urgently needed to provide additional evidence of kidney impairment in COVID-19 patients.

In the current study, we enrolled 178 patients with confirmed SARS-CoV-2 infection including 52 cases admitted to ICU and 126 cases not. We found no patient exhibited a rise in Scr or Cystatin C and occurrence of AKI both in ICU group and non-ICU group. On the contrary, 54.2% (45/83) patients who performed urine routine tests (48.9% [87/178]) presented abnormal urinalysis, which was featured by proteinuria, hematuria and leucocyturia. These results reveal that renal impairment are common both in severe and non-severe COVID-19 patients, which is manifested by abnormal urine routine test rather than elevated level of Scr and AKI. Meanwhile, our study indicates that urinalysis is a better tool to screen kidney impairment.

These two seemingly opposite results based on blood chemistry analysis and urinalysis respectively arouse our interest in exploring the underlying reasons. Firstly, although it’s believed that SARS-CoV-2 infection causes kidney impairment, clinical symptoms were not obvious due to the powerful compensatory function of kidney. It’s well known that serum levels of Scr and BUN will exceed the normal range only when more than 50% of kidney function has been lost. The extent of SARS-CoV-2 infection caused kidney impairment may be not enough to result in elevated plasma markers related renal function. In other words, “kidney impairment” doesn’t lead to “kidney dysfunction” to a certain extent. Secondly, the primary damage is restricted in renal tubules rather than glomerulus **(Figure 4)**. Several investigations revealed that SARS-CoV-2 gain entry into host cells by binding to ACE2 receptor on the host cell surface as the SARS-CoV does. Previous study of ACE2 immunostaining revealed that the mesangium and glomerular endothelium were negative for ACE2 and the distal tubules and collecting ducts showed weak cytoplasmic staining, whereas abundant staining was seen in the brush border of proximal tubular cells(22). Another two studies conducted by Xu et al and Qi’s group pointed that proximal straight tubule cells were potential host cells targeted by SARS-CoV-2 by scRNA-seq analysis(12, 23). Indeed, such speculation has already been verified by Diao’s group who reported immunohistochemistry of SARS-CoV-2 NP antigen in kidney specimens of six perished cases and found that NP expression was restricted in kidney tubules and absent in the glomerulus. Histopathological examination by H&E staining in kidney specimens has proved acute tubular necrosis, luminal brush border sloughing and vacuole degeneration(24). Furthermore, another study on organ distribution of SARS-CoV in patients who died of SARS reported that SARS-CoV was detected in epithelial cells of distal convoluted renal tubules(25). Meanwhile, the pathological findings of kidney form autopsy in seven SARS subjects pinpointed that glomerular pathology was not observed in the kidneys, but acute tubular necrosis of varying degrees was found in all seven renal specimens(26).

**Figure 4.**
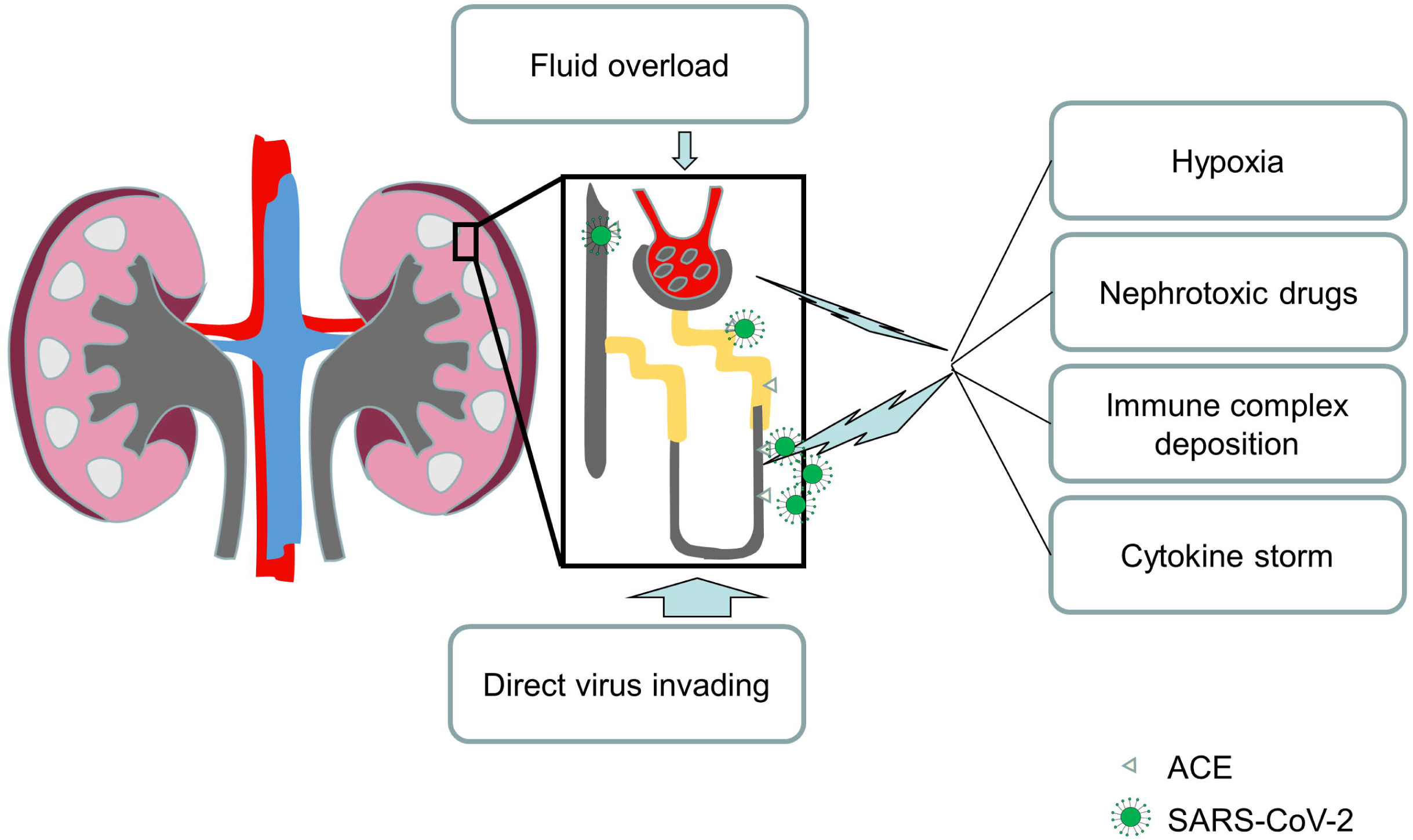
Potential mechanisms of kidney impairment in COVID-19patients. SARS-CoV-2 can directly invade renal tubular epithelial cells via ACE2, a virus specific receptor that mainly expressed in proximal tubular cells, which potentially leading to host cells damage and death. At the same time, nephrotoxic drugs, hypoxia and infectious caused extensive immune activation, featured by cytokine storm and immune complex formation, were also factors related to tubular cells impairment. The impairment caused compromise of tubular integrity may be major reason that the occurrence of abnormal urinalysis. In addition, although no significant signs of kidney dysfunction and acute kidney injury, characterized by increased Scr, BUN and cystatin C, many known factors may provoke injury to the glomeruli, such as inflammation, nephrotoxic drugs and fluid overload.

So far, many parameters and models were established to predict the change of condition among COVID-19 patients. In order to evaluate whether urinalysis is useful to predict the disease severity, we then divided the patients into two groups based on their results of urine routine test on admission, named abnormal urinalysis group (AU) and normal urinalysis group (NU). Compared to control patients, patients in AU group presented no difference in BUN, Scr and eGFR, but had elevated cystatin C. In addition, we found that patients with abnormal urinalysis usually had more severe laboratory parameters, such as higher liver and heart injury index, including ALT, AST, ALP, LDH, HBDH, GGT and LAP, inflammation related markers, including CRP, IL-6, ESR, SAA and serum ferritin, blood routine indicator including WBC, neutrophils and lymphocytes, as well as coagulation related indicator including FIB and D-dimer. This analysis revealed that the patients with kidney impairment were generally in worse condition, including multiple organs or tissues injuries (liver, heart), inflammation and hypercoagulability state, which is probably because of low level of immunity and high viral load. In addition, our study indicates that urinalysis, a cheap, fast and easy way to perform, is an excellent method to predict the severe condition of COVID-19 patients.

Admittedly, there are several issues need to be addressed in this study. Firstly, we have no pathological evidence to confirm our speculation due to lack of specimen from biopsy or autopsy, such as H&E and immunostaining. Secondly, the dynamic analysis of urinalysis was not performed in COVID-19 patients due to many patients only have one urine routine test on admission. Finally, a larger sample size should be enrolled in our study. We will attempt to address these questions in our future research.

In summary, we provided the evidence that kidney impairment is common among COVID-19 patients, and its clinical manifestation is abnormal urinalysis, indicating that urine routine test is a better indicator to unveil potential kidney impairment than blood chemistry test. Furthermore, our results suggested that urinalysis is a useful tool to predict disease severity. Thus, we call for front-line healthcare workers pay more attention on kidney impairment in patients with SARS-CoV-2 infection and routinely monitor urinalysis to judge potential kidney impairment and evaluate disease severity.

## Data Availability

The data used to support the findings of this study are available from the corresponding author upon request.

## Contributors

DH and HF designed the study. HZ, ZZ prepared figures and tables, and wrote the paper. ML, YD, WG, LL, ZK, TY, CT, YG, RQ collected the data and analyzed the data. JL revised the paper. HW, SL and SL participated in the discussion of this study and give suggestions on formulation of the manuscript.

## Acknowledgments

This study was funded by the grants from the project of Thousand Youth Talents for D.H.; and from the China National Natural Science Foundation (Nos. 31770983 and 81974249 to D.H., No. 81601747 to S.L.). The authors thank all members of Wuhan Union Hospital for helpful suggestions and discussions in our study.

